# Metronidazole and *Giardia*: *in vitro* viability assay under microaerophilic conditions indicates a multifactorial basis for metronidazole treatment failure

**DOI:** 10.1101/2025.02.26.25322590

**Authors:** Eva Nohýnková, Aneta Perglerová, Pavla Tůmová

## Abstract

We developed a simple 24-hour viability assay to evaluate the tolerance of the parasite *Giardia intestinalis* to metronidazole (MTZ) *in vitro*. Our findings emphasize the importance of test conditions and evaluation parameters. Using this assay with minimum lethal concentration (MLC) as an evaluation parameter, we identified two isolates that are naturally tolerant to MTZ. Natural MTZ tolerance was detected only under microaerophilic conditions, as both isolates were susceptible to MTZ under anaerobic conditions. These isolates were derived from stool samples of patients with MTZ-refractory giardiasis, suggesting that the parasite tolerance was responsible for the failure of MTZ treatment. However, two other isolates from patients with MTZ-refractory giardiasis were susceptible under both test conditions, indicating that MTZ-refractory giardiasis likely has a multifactorial cause.

**HIGHLIGHTS:**

- *In vitro* test conditions are significant to assess metronidazole tolerance in *Giardia*
- *In vitro* test should be done under both anaerobic and microaerophilic conditions
- MLC, but not IC_50_, is a valuable parameter for evaluating MTZ tolerance
- Failure to treat giardiasis with metronidazole probably has a multifactorial cause

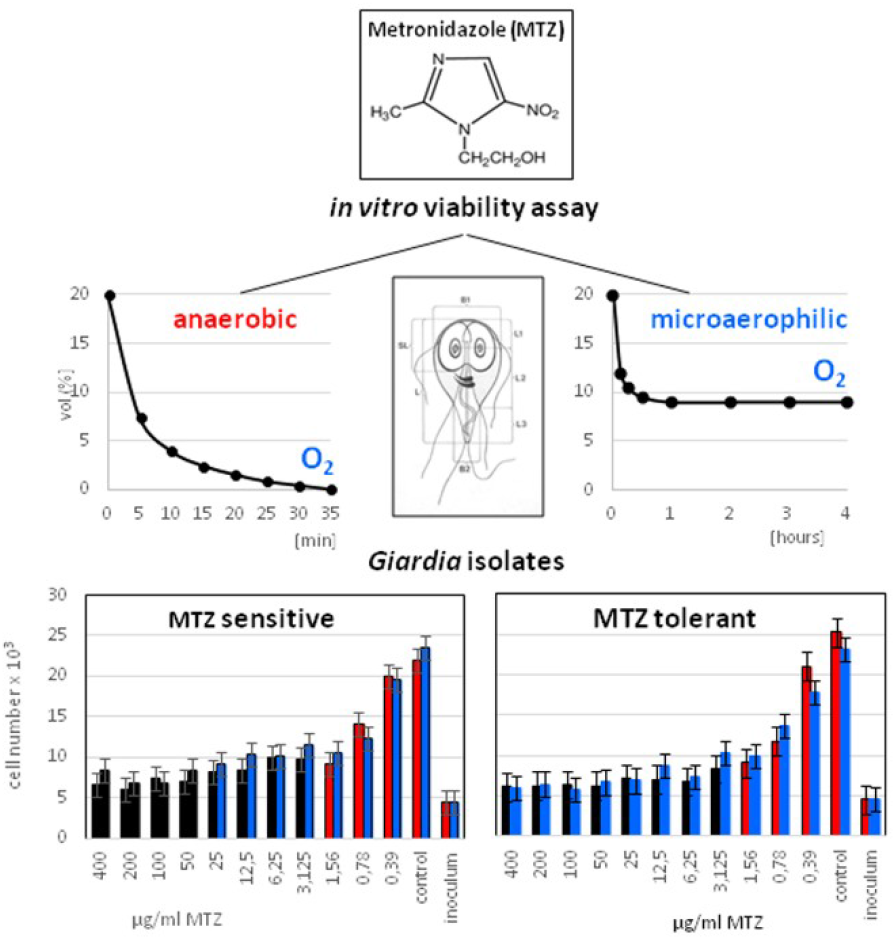

## 1. Introduction

Giardiasis is a parasitosis affecting the small intestine of humans and other mammals worldwide. In humans, the clinical picture of giardiasis varies significantly from asymptomatic infection to diarrhea with steatorrhea, flatulence and abdominal discomfort. Malabsorption syndrome in young children or postinfectious sequelae such as chronic fatigue or irritable bowel syndrome in adults are well-defined complications of chronic giardiasis (Litleskare et al., 2018). The infection caused by the pathogenic flagellate *Giardia intestinalis* is usually treatable with metronidazole (MTZ), a drug of choice for the treatment of anaerobic infections of both bacterial and protozoan origin. However, in recent years, the number of giardiasis cases refractory to MTZ treatment has increased (Lalle and Hanevik, 2018; Carter et al., 2018; Stejskal et al., 2015). It is therefore paradoxical that according to available published data, no *Giardia* isolate with proven natural MTZ resistance has yet been isolated *in vitro*, and there is only one report that mentions the resistance of clinical isolates to MTZ in *in vivo* experiments in a mouse model (Lemée et al., 2000).

MTZ, similar to other 5-nitroimidazoles, is a prodrug, meaning that it must be partly metabolized within a cell to generate toxic reductive species that have an impact on various pathogen biomolecules and metabolic pathways (Dingstag and Hunter, 2018; Uzlíková and Nohýnková, 2014). The microbicidal effect of 5-nitroimidazole drugs is based on a serial reduction of a nitro group at position 5 on the imidazole ring into toxic nitro- and nitroso-radicals via electrons generated in/by anaerobic metabolism. In contrast to bacteria and other protozoan anaerobes infecting humans, namely *Trichomonas vaginalis* and *Entamoeba histolytica*, in which the intracellular reduction of MTZ is primarily mediated via electrons generated by pyruvate ferredoxin oxidoreductase (PFOR), additional enzymes are likely to reduce MTZ in *Giardia* (reviewed in Leitsch, 2015; Leitsch et al., 2018; Argüello-García et al., 2020). When present, intracellular oxygen, which is a strong electron acceptor, competes with the drug for electrons generated via the anaerobe’s metabolism. Therefore, the sensitivity of any anaerobe, including *Giardia*, to MTZ is significantly affected by the concentration of oxygen present in the environment (e.g., the intestinal lumen) and the oxygen-detoxifying potential within the pathogen cell. In the small intestine where *Giardia* resides and multiplies during infection, a steep oxygen gradient along the crypt-villus axis has been documented (reviewed in Zheng et al., 2015; Zeitouni et al., 2016). In the crypt, the physiological oxygen concentration is approximately 8% (partial pressure (pO_2_) of 59 mm Hg), while it is approximately 3% (pO_2_ of 22 mm Hg) at the tip of the villus (Zeitouni et al., 2016; Singhal and Shah, 2020). Moreover, the small intestine is subject to pronounced fluctuations in oxygen concentration associated with periodic ingestion/digestion/absorption of nutrients. *Giardia* trophozoites, a pathogenic stage of the parasite, typically reversibly adhere along the villi of the duodenum–jejunum epithelium by means of a *Giardia*-specific attachment organelle, the ventral disc. *Giardia* thus inhabits a specific microaerophilic environment, with a low total microbial density estimated at 10^3^ – 10^5^ cells/g, reflecting unfavorable conditions due to nutrient digestion (digestive enzymes, pancreatic, gastric and bile acids, etc.) (Ruigrok et al., 2023). Additionally, a limited number of bacterial pathogens colonize this part of the intestine. One of them is the gram-negative bacterium *Helicobacter pylori* and, likely, the closely related bacterium *Campylobacter jejuni*, a leading cause of bacterial enteritis in humans (Burnham and Hendrixson, 2018). Both of these pathogens are microaerophils with some degree of oxygen tolerance. We therefore tested the *in vitro* sensitivity of *Giardia* to MTZ under anaerobic (<1% oxygen) and microaerophilic (>5% oxygen) conditions used for *in vitro* propagation of the facultative anaerobe/microaerophil *H. pylori* (Blanchard and Nedrud, 2012) to determine the effect of physiological oxygen concentrations on *Giardia* survival under MTZ pressure. In these tests, we applied our collection of *Giardia* laboratory isolates derived from giardiasis patients with mostly known responses to MTZ treatment. Similar approach was recently published by Starynets et al. (2025), who found increased expression of antioxidant enzymes in two laboratory strains growing under microaerophilic conditions.

## 2. Materials and methods

### 2.1 Giardia isolates

For the MTZ sensitivity tests, 18 clinical and 2 reference isolates of *G. intestinalis* were used. A list of isolates is given in Tab. 1. Some characteristics of the isolates can be found in Lecova et al. (2018). Seven of these isolates were derived from patients with MTZ-refractory giardiasis (one of them, the NAX isolate, was from a patient who exhibited treatment failure with multiple drugs), and 11 isolates were from giardiasis patients who responded well to routine MTZ treatment as verified microscopically 4 weeks after the end of the treatment. Cysts were purified from stool using sucrose gradient and in most cases the simplified excystation protocol of Feely was used (see Kulda and Nohýnková, 1995, p. 317, Table 3.6) with the exception of the ZED isolate, where the excystation protocol of Meng et al (1996) was used. Isolates HP-1, a Prague line of Portland-1, ATCC 3088, provided by E.A. Meyer (Oregon Health Science University) and WBc6 (a clone of the WB isolate, ATCC 30957) were used as reference isolates. Before testing, the isolates were maintained anaerobically in 7.5 ml of TYI-S-33 culture medium, pH 6.8, with final concentrations of 0.2 ‰ ferric ammonium citrate, 0.1% bovine bile and 10% bovine serum in borosilicate glass culture tubes with screw caps (13x100 mm; Bellco). Similarly, freshly prepared medium was used for *in vitro* tests. All the procedures in the study involving human material and data were performed in accordance with the World Medical Association’s Declaration of Helsinki – Ethical Principles for Medical Research Involving Human Subjects (WMA, 2013). The isolate IDs were not known to anyone outside the research group.

### 2.2 MTZ tests

*In vitro* tests were performed in sterile flat bottom 96-well culture microplates (Costar, Corning) in a total volume of 200 µl per well. A stock solution of MTZ (Sigma) was prepared immediately before use by autoclaving 16 mg of MTZ in 1 ml of double-distilled H_2_O, to which 4 ml of TYI-S-33 culture medium was added after cooling to produce a solution of 3.2 mg of MTZ/ml.

Each test was performed as follows: Into each well of a microplate, 50 µl of TYI-S-33 medium was dispensed using a stepper. To each well of the second column (wells A2 through H2), 50 µl of the MTZ stock solution was added, and the solution was serially diluted using an 8-channel micropipette by transferring 50 µl of a solution from the wells in the second column to the wells in the third column and so on (a twofold dilution). Between each transfer, the solution in the well was mixed with the micropipette. After diluting MTZ, suspensions of the tested isolates were prepared. For each isolate, 8 ml of a culture (with a *Giardia* monolayer) and one autoclavable reagent reservoir (60 ml; Thermo Fischer Scientific) were employed per test. Immediately before use, a sterile reagent reservoir wrapped with aluminum foil was cooled in a freezer for 5 minutes. In parallel, a culture tube with the tested isolate was incubated on ice for 5 minutes. Then, the culture was thoroughly resuspended to detach the *Giardia* cells from the tube walls, and the cell suspension was poured from the tube into the precooled reservoir (to prevent *Giardia* cells from adhering) and inoculated immediately. Then, 150 µl of the suspension was added to each well of one row using a 12-channel micropipette. In this way, in each row of the microplate, there was no MTZ in the first well (a control well), a concentration of 400 µg/ml MTZ (2.3 mM; the highest concentration tested) was present in the second well, and 0.39 µg/ml MTZ (2.3 µM; the lowest concentration tested) was present in the twelfth well. After inoculation of all tested isolates, the microplate was shaken with a microplate shaker for 30 seconds and incubated in an appropriate gas mixture in a plastic pouch at 37°C for 24 hours. In each experiment, two microplates were prepared in parallel for testing under anaerobic and microaerophilic conditions. For each isolate, two rows per microplate for anaerobic and microaerophilic conditions were used in each experiment. At least three independent experiments were performed for each isolate. To calculate the inoculum, 100 µl of the initial suspensions was used.

For tests under anaerobic conditions, AnaeroGen compact (Oxoid), an anaerobic atmosphere generation system, was used. For tests under microaerophilic conditions, a CampyGen compact (Oxoid), a system generating atmosphere for the growth of microaerophils, was used. Both systems consist of a W-Zip plastic pouch and a paper gas generating sachet. AnaeroGen produces an atmosphere consisting of < 1% O_2_ and 9-13% CO_2_. CampyGen produces a gas mixture consisting of 8-9% O_2_ and 7-8% CO_2_ (Thermo Scientific^TM^ Oxoid^TM^). In both systems, N_2_ but not H_2_ complements the gas mixture.

### 2.3 Test evaluation

Tests were evaluated using two parameters: minimum lethal concentration (MLC) and 50% inhibitory concentration (IC_50_). The MLC was defined according to Upcroft et al. (1999) as the lowest concentration of the drug at which no motile cells were detected by microscopic examination. The MLC was evaluated well by well using an inverted microscope (Olympus) immediately after the end of the 24-hour incubation.

The IC_50_ was defined as the concentration of the drug required to achieve a cell number of 50% of that of the control and was evaluated as follows. Immediately after MLC evaluation, the cells were fixed with formalin for counting. Briefly, 100 µl of 1% formalin/PBS was added to each microplate well containing 200 µl of the tested culture, the microplate was then briefly shaken with a microplate shaker to mix the fixative with the well contents, and the microplate with a lid was placed in a refrigerator overnight. In this way, each *Giardia* cell, live or dead, was fixed and detached from the well bottom. The next day, the contents of each well were manually mixed using a standardized procedure with a micropipette (the micropipette was set to 250 µl; the tip was filled and emptied 10 times), and the cell numbers were determined microscopically by using a Bürker counting chamber.

## 3. Results

### 3.1 Drug sensitivity evaluation

#### 3.1.1 Minimal lethal concentration (MLC)

Under anaerobic conditions, none of the 18 clinical isolates exhibited decreased sensitivity to MTZ. The MLC ranged between 3.13 - 12.5 μg/ml MTZ and was similar for all tested isolates as well as for the reference strains (Tab. 1).

**Table 1.** A list of isolates of *Giardia intestinalis* tested *in vitro* for tolerance to metronidazole (MTZ) under anaerobic and microaerophilic conditions. 24hrs exposure. Evaluating parameters: MLC and IC_50_.

| isolate ID | clinical resistance | MLC anaerobic conditions (µg/ml) | MLC microaerophilic conditions (µg/ml) | IC<sub>50</sub> anaerobic conditions (µg/ml) | IC<sub>50</sub> microaerophilic conditions (µg/ml) |
| --- | --- | --- | --- | --- | --- |
| 2 | NO | 12.5 | 50 | nd | nd |
| 8 | NO | 6.25 | 100 | nd | nd |
| 9 | NO | 6.25 - 12.5 | 100 - 200 | nd | nd |
| 12 | NO | 3.13 | 50 | 0.78 | 0.78 |
| 13 | NO | 6.25 | 100 | nd | nd |
| 18 | NO | 6.25 – 12.5 | 25 - 50 | nd | nd |
| 19 | NO | 6.25 – 12.5 | 50 | nd | nd |
| 25 | NO | 6.25 | 50 | nd | nd |
| 28 | NO | 3.13 | 50 | 1.56 | 1.56 |
| 34 | NO | 3.13 | 100 | nd | nd |
| 49 | NO | 6.25 | 12.5 | nd | nd |
| 16 | YES | 3.13 | 50 | nd | nd |
| 23 | YES | 12.5 | 100 | nd | nd |
| 24 | YES | 18.3 – 12.5 | 100 - 200 | 0.78 – 1.56 | 0.78 – 1.56 |
| BER-1 | YES | 18.3 - 6.25 | > 400 | 0.78 | 0.78 – 1.56 |
| HH | YES | 3.13 | 12.5 | nd | nd |
| ZED | YES | 6.25 | > 400 | nd | nd |
| NAX | YES | 3.13 | 25 | nd | nd |
| WBc6 | r | 3.13 | 50 | nd | nd |
| HP-1 | r | 3.13 | 50 | nd | nd |

nd – not done r – reference isolate

Under microaerophilic conditions, i.e., in the presence of > 5% O_2_, the isolates exhibited significantly greater tolerances to MTZ between 12.5 and 400 μg/ml. Two clinical isolates (BER-1 and ZED) were highly tolerant to MTZ, with an MLC >400 μg/ml MTZ (Tab. 2), and were considered MTZ resistant *in vitro*. Another two isolates (9, 24) showed increased tolerance to MTZ, with MLCs varying between 100 and 200 μg/ml MTZ, and were considered MTZ tolerant. Fourteen other isolates with sensitivities to MTZ in the MLC range between 12.5 and 100 μg/ml were considered MTZ sensitive. Interestingly, two of these isolates that were most susceptible to MTZ (MLC 12.5 μg/ml) were previously genotyped as assemblage B (Lecova et al., 2018, 2019), while twelve isolates with higher MTZ tolerance (MLC 50-100 μg/ml) belonged to assemblage A (Lecova et al., 2018).

**Table 2.** *In vitro* isolates from four cases of giardiasis unresponsive to treatment with standard dose metronidazole.

| isolate ID | drug | treatment history dosage |
| --- | --- | --- |
| BER-1 | MTZ | 250mg bid/10d |
|  | MTZ | 250mg bid/10d |
|  | OZ | 500mg bid/6d |
|  | MTZ | 250mg bid/10d |
|  | ALB | 400mg bid/7d |
|  | MTZ+ALB | 250mg tid + 400mg bid/7d |
| ZED | MTZ | 250mg bid/7d |
|  | MTZ | 500mg bid/7d |
|  | OZ | 500mg bid/6d |
| HH | MTZ | 250mg bid/7d |
|  | MTZ | 250mg tid/7d |
| NAX | MTZ | 250mg bid/10d |
|  | MTZ | 250mg tid/10d |
|  | PMC | 3-2-3<sup>1</sup>/5d |
|  | MTZ+ALB | na |
|  | na | na |
|  | na | na |
|  | NT | na |
|  | OZ | na |
|  | NT | 1000mg bid/2d + 500mg bid/1d |

MTZ – metronidazole; OZ – ornidazole; ALB – albendazole; PMC – paromomycin; NT – nitazoxanide; na – not available
<sup>1)</sup> 2g/d divided q8hr

#### 3.1.2 Inhibitory concentration 50% (IC_50_)

To determine the utility of the IC_50_ as a parameter for evaluating the *in vitro* sensitivity of *Giardia* isolates to MTZ, we selected four clinical isolates (12, 24, 28, BER-1) representing samples with different tolerances to MTZ detected by MLC evaluation and from patients with giardiasis with different responses to MTZ treatment (Tab. 1). Unlike the MLC, the IC_50_ showed no variability in the MTZ sensitivity. Under both anaerobic and microaerophilic conditions, there was no difference in the MTZ concentration needed to inhibit the growth of *Giardia* trophozoites to 50% of the control cell growth after 24 h of exposure. In both gas mixtures, the IC_50_ was 0.78-1.56 µg/ml MTZ for all four tested isolates (Tab. 1 and Fig. 1).

**Figure 1.**
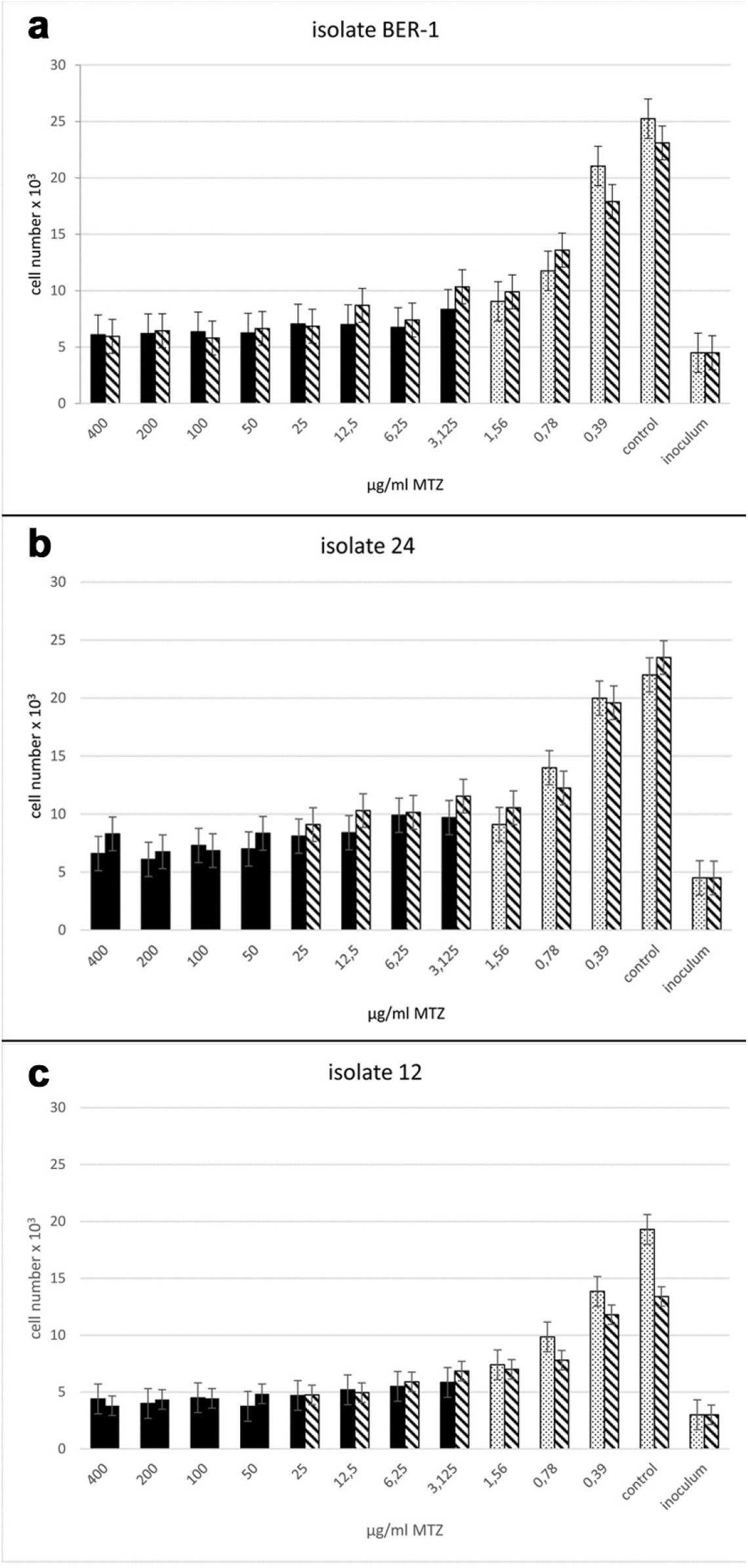
Inhibitory concentration 50 (IC_50_) and minimum lethal concentration (MLC) of metronidazole (MTZ) for three *Giardia* isolates after 24-hour *in vitro* exposure to MTZ under microaerophilic and anaerobic test conditions. Anaerobic conditions (dotted columns); microaerophilic conditions (hatched columns). In each isolate, cell numbers representing 50% inhibition of control cells growth (control column), i.e. IC_50_, were between 0.78-1.56 µg/ml MTZ independent of the gas mixture (a-c). Black columns represent MTZ concentrations at which no live/motile cells were observed by microscopic examination, meaning that the first MTZ concentration without live cells represents the minimum lethal concentration (MLC) of MTZ for the isolate under the given conditions. Under anaerobic conditions (left in each pair of columns), the MLC was 3.125 µg/ml MTZ for all three isolates shown. Under microaerophilic conditions (right in each pair of columns), the MLC was 50 µg/ml MTZ for isolates 24 and 12 (b, c), and more than 400 µg/ml MTZ for isolate BER-1 (a).

### 3.2 MTZ tolerance of *Giardia in vitro* vs. MTZ-refractory giardiasis

Our collection included four *Giardia* isolates derived from patients with MTZ-refractory giardiasis with a well-documented history of treatment failure (Tab. 2). Of these, two clinical isolates, BER-1 and ZED, showed high and stable tolerance to MTZ, as assessed by determining the MLC under microaerophilic conditions (>400 μg/ml MTZ). Under the same conditions, two other isolates, HH and NAX, from the same group of four patients with well-documented cases of MTZ treatment failure did not show any *in vitro* MTZ tolerance, with MLC concentrations of 12.5 and 25 μg/ml MTZ (Tab. 2).

## 4. Discussion

### 4.1 Drug sensitivity evaluation

To our knowledge, no laboratory isolate of *G. intestinalis* naturally resistant to MTZ has been derived from a patient, although giardiasis treatment failure is common. Thus, it is still unclear to what extent clinical unresponsiveness to MTZ is due to MTZ-resistant parasites or even whether such parasites exist in nature. As there are still no molecular markers that can be used to demonstrate resistance/sensitivity to MTZ in *Giardia* (Müller et al., 2019), traditional *in vitro* assays remain the first option for this purpose. Whether the procedures/methods used thus far can provide relevant information is unclear.

Regarding *in vitro* testing, several older papers have been devoted to this topic (Tab. S1) (Boreham et al., 1984; McIntyre et al., 1986; Upcroft et al., 1990; Majewska et al., 1991; Favennec et al., 1992; Farbey et al., 1997; Adagu et al., 2002). Unfortunately, the conditions used in these experiments, including the system (experimental volume), time of MTZ exposure (3 to 72 h), assay/procedure (^3^H-thymidine uptake, adherence test, clonal growth, viability), and parameters evaluated (MIC, IC_50_, ID_50_, and MLC), were heterogeneous, making it very difficult to compare their results (Tab. S1). Moreover, most of the experiments were performed without a controlled/defined gas atmosphere.

Our results show the crucial importance of experimental design for the *in vitro* detection of MTZ tolerance in *Giardia* in terms of both the evaluated parameters and the gas atmosphere. The IC_50_ represents one of the most commonly used parameters for testing *in vitro* drug efficacy against microbes, incl. *Giardia*. However, we have shown that the IC_50_ defined as growth inhibition to 50% of that of a control cell population is unusable when assessing MTZ sensitivity in *Giardia*. The MTZ concentrations required to inhibit the growth of four tested isolates to 50% of the growth of untreated cells were the same regardless of the gas atmosphere, MLC or clinical resistance of a particular isolate. In contrast, the MLC proved to be a valuable parameter for the evaluation of *Giardia* MTZ susceptibility. However, as shown in our experiments, adjusting the incubation atmosphere to the appropriate oxygen concentration is essential.

When using systems for generating an anaerobic atmosphere, e.g., AnaeroGen (Oxoid) or the GasPak EZ Anaerobe System (Becton Dickinson), less than 1% O_2_ is present in the gas mixture. According to published data, in several cases where the MTZ sensitivity of *Giardia* isolates has been tested *in vitro* under a controlled gas atmosphere, anaerobic conditions were used (Ellis et al., 1993). Under these conditions, all of the isolates, except the *in vitro*-derived MTZ-tolerant line BRIS/83/HEPU/106-2ID_10_ (Boreham et al., 1988), were susceptible to the drug. This was also the case for all our isolates when tested anaerobically. There were minimal differences among the isolates at the three lowest concentrations of MTZ, so the isolates were clearly susceptible to the drug. It seems that anaerobically resistant *Giardia* isolates are rare, if present, in nature. One reason could be that similar to that for *Trichomonas vaginalis* (Kulda et al., 1993), anaerobic resistance is likely achieved after long-term permanent exposure of *Giardia* to nonlethal doses of the drug. In this way, several MTZ-resistant laboratory lines commonly used to study MTZ resistance in *Giardia*, namely, BRIS/83/HEPU/106-2ID_10_, WB-C6-NTZ/METrC4 and WB-C6-METrC5, were generated *in vitro* (Boreham et al., 1988; Muller et al., 2007). Additionally, experiments with these lines showed that anaerobic resistance can be lost even during the *in vitro* encystation/excystation cycle (Muller et al., 2008). They likely exhibit unstable MTZ resistance because they must be maintained under permanent MTZ exposure/pressure; otherwise, resistance disappears (Boreham et al., 1988; Muller et al., 2018). Whether this phenomenon might also occur naturally is unknown. Ansell et al. (2017) speculated that if such isolates are generated from patient-excreted cysts via excystation and further propagated *in vitro* in drug-free culture medium for some period prior to testing, they may lose their initial/original MTZ tolerance. However, no experimental data supporting this speculation are available.

### 4.2 MTZ-refractory giardiasis vs. *Giardia* MTZ tolerance *in vitro*

Of our seven isolates from MTZ-refractory patients, four were isolated from patients with well-documented treatment. Interestingly, we found that under microaerophilic conditions, two strains (BER-1 and ZED) were highly tolerant to MTZ, while the other two (HH and NAX) were sensitive. Moreover, the MTZ tolerance of the BER-1 and ZED isolates was stable and independent of isolate maintenance/manipulation (duration of cultivation, cryopreservation, etc.). They were first tested for *in vitro* sensitivity to MTZ nearly twenty years ago (Nohynkova et al., 2004) and several times since, with the same results as in this paper (Nohynkova unpublished). Long-lasting MTZ tolerance can be regarded as a natural characteristic of an isolate that manifests only in the presence of oxygen. Thus, this type of MTZ resistance likely corresponds to the so-called aerobic type described previously in human trichomonads (Tachezy et al., 1993), but in *Giardia*, the molecular background of the “aerobic type” of MTZ tolerance is unknown. Because these two isolates were derived from patients with MTZ-refractory giardiasis, in these particular cases, the MTZ tolerance of the pathogen may be responsible for treatment failure, which is clearly attributable to the parasite. On the other hand, in two other cases of giardiasis, the failure of MTZ treatment cannot be attributed to the parasite, as both isolates (HH and NAX) showed no tolerance to MTZ *in vitro*. Thus, it seems clear that MTZ-refractory giardiasis is likely to have a multifactorial basis, highlighting the complexity of MTZ treatment failure.

## Data Availability

All data produced in the present work are contained in the manuscript

## Declaration of sample IDs

The authors declare that the isolate IDs are not known to anyone outside the research group.

## Acknowledgements

This work was partly supported by the Czech Health Research Council (grant No. NU23-05-00441).

