## Supplemental Table 1 for "Metronidazole and *Giardia*: *in vitro* viability assay under microaerophilic conditions indicates a multifactorial basis for metronidazole treatment failure"

**Table S1. Published data on metronidazole sensitivity tests used for *in vitro* testing of *Giardia* isolates**

| origin | *In vitro* assay | gas atmosphere | time of exposure (hrs) | evaluating parameter | isolate | sensitivity dose (µM) [1)] |
|---|---|---|---|---|---|---|
| Gillin and Diamond 1981 | clonal growth | nd | 5 – 7 days | $IC_{50}$ | likely PORTLAND 1 | **11.66** |
| | | | | MLC | | **11.66 – 23.32** |
| Smith et al. 1982 | clonal growth | nd | 5 – 7 days | $IC_{50}$ [2)] | WB | **9.33** |
| | | | | MLC [3)] | PORTLAND 1 | **11.08** |
| | | | | | LT | **9.33** |
| | | | | | RS | **5.83** |
| | | | | | WB | **21.86** |
| | | | | | PORTLAND 1 | **21.86** |
| | | | | | LT | **21.86** |
| | | | | | RS | **10.96** |
| Boreham et al. 1984 | $^{3}H$-thymidine incorporation | nd | 24 | $ID_{50}$ [4)] | BRIS/82/HEPU/41 | 2.14 |
| | | | | | BRIS/83/HEPU/99 | 1.61 |
| | | | | | BRIS/83/HEPU/106 | 0.91 |
| | | | | | BRIS/83/HEPU/120 | 1.08 |
| | | | | | BRIS/83/HEPU/136 | 2.21 |
| | | | | | PORTLAND 1 | 1.79 |
| Boreham et al. 1985 | $^{3}H$-thymidine incorporation | nd | 24 | $ID_{50}$ [4)] | BRIS/83/HEPU/106 | 0.87 |
| | | | | | BRIS/83/HEPU/99 | 1.08 |
| McIntyre et al. 1986 | $^{3}H$-thymidine incorporation | nd | 24 | $ID_{50}$ [4)] | 13 isolates | 0.99 ± 0.15 |
| Smith et al. 1988 | $^{3}H$-thymidine incorporation [5)] | nd | 24 | $ID_{50}$ [4)] | BRIS/85/HEPU/436 | 0.45 |
| | | | | | BRIS/84/HEPU/343 | 0.60 |
| | | | | | BRIS/83/HEPU/141 | 0.61 |
| | | | | | BRIS/85/HEPU/452 | 0.66 |
| | | | | | BRIS/83/HEPU/210 | 0.68 |
| | | | | | BRIS/83/HEPU/153 | 0.71 |
| | | | | | BRIS/83/HEPU/120 | 0.78 |
| | | | | | BRIS/85/HEPU/449 | 0.83 |
| | | | | | BRIS/83/HEPU/106 | 0.86 |
| | | | | | BRIS/83/HEPU/161 | 0.91 |

| | | | | | | |
|---|---|---|---|---|---|---|
| | | | | | BRIS/84/HEPU/353 | 0.92 |
| | | | | | BRIS/85/HEPU/497 | 1.04 |
| | | | | | BRIS/83/HEPU/99 | 1.07 |
| | | | | | BRIS/86/HEPU/592 | 1.15 |
| | | | | | BRIS/82/HEPU/41 | 1.77 |
| | | | | | PORTLAND 1 | 1.79 |
| | | | | | BRIS/83/HEPU/136 | 2.21 |
| | | | | | BRIS/86/HEPU/567 | 4.48 |
| Upcroft et al. 1990 | $^3$H-thymidine incorporation | nd | 24 | ID$_{50}$ | Ad113 | 0.55 |
| | | | | | BRIS/89/HEPU/1003 | 0.65 |
| | | | | | BRIS/83/HEPU/106 | 0.77 |
| | | | | | BRIS/83/HEPU/106-2ID$_{10}$ | 8.50 |
| Ellis et al. 1993 | $^3$H-thymidine incorporation | nd | 24 | ID$_{50}$ | BRIS/83/HEPU/120 | 0.98 |
| | | | | | OAS1 (sheep isolate) | 0.75 |
| | | | | | WB1B | 1.15 |
| Adagu et al. 2002 | $^3$H-thymidine incorporation | 3% $O_2$/4% $CO_2$/93% $N_2$ | 48 | ID$_{50}$ | JKH-1 | 15.42 ± 4.3 |
| | | | | | EBE | 5.95 ± 5.8 |
| | | | | | VNB1 | 5.72 ± 3.4 |
| | | | | | VNB5 | 5.20 ± 3.5 |
| | | | | | EBC | 4.55 ± 2.2 |
| | | | | | VNB2 | 3.21 ± 1.2 |
| Favennec et al. 1992 | adherence test [6)] | nd | | | | **8.77** |
| | | likely anaerobic | 18 | ID$_{50}$ | PARIS/86/LCF/3 | |
| | growth inhibition | | | | | **6.72** |
| Farbey et al. 1995 | adherence test [7)] | nd | 4 | IC$_{50}$ | 29 isolates | 0.009 – 153.6 |
| | | | | | P1C10 | 0.31 |
| Abboud et al. 2001 | adherence test [8)] | nd | na (18) | IC$_{50}$ | isolate from HIV+ patient | **>1166** |
| Cruz et al. 2003 | adherence test | nd | na (4) | IC$_{50}$ | ACPT 98004 | 3.38 |
| | | | | | ACPT 98005 | 4.25 |
| | | | | | ACPT 98006 | 7.49 |
| | | | | | ACPT 98013 | 5.52 |
| | | | | | ACPT 98012 | 2.38 |
| | | | | | ACPT 98014 | 3.60 |

| | | | | | | |
|---|---|---|---|---|---|---|
| | | | | | ACPT 98018 | 3.84 |
| | | | | | ACPT 98019 | 10.09 |
| | | | | | ACPT 98020 | 11.50 |
| | | | | | ACPT 98021 | 4.28 |
| | | | | | ACCA 98007 | 3.01 |
| | | | | | ACCA 98010 | 4.92 |
| | | | | | ACPT 98008 | 3.33 |
| | | | | | ACPT 98009 | 3.34 |
| | | | | | ACPT 98011 | 4.46 |
| | | | | | ACPA 99015 | 2.52 |
| | | | | | ACPT 99017 | 7.18 |
| | | | | | ACPT 99016 | 3.41 |
| | | | | | ATCC 30888 (PORTLAND 1) | 3.10 |
| | | | | | ATCC 30957 (WB) | 4.27 |
| Cedilo-Rivera and Munoz 1992 | growth inhibition | nd | 48 | $IC_{50}$ [9)] | PORTLAND 1 | **1.23** |
| | | | | MLC [10)] | | **5.85 – 29.25** |
| Upcroft et al. 1999 | viability | nd | 72 | MLC [11)] | BRIS/83/HEPU/106 | 100 |
| | | | | | BRIS/83/HEPU/106-2ID$_{10}$ | >500 |
| | | | | | BRIS/89/HEPU/1279 | 50 |
| | | | | | WB1BM3 | >500 |
| Céu Sousa and Poiares-da-Silva 1999 | oxygen uptake | likely anaerobic | 3 | | ATCC 30957 (WB) | 330 |
| Ansell et al. 2017 | viability [12)] | anaerobic <0.7% $O_2$ ~10% $CO_2$ | 48 | $IC_{50}$ | WB1B | 8.28 |
| | | | | | BRIS/83/HEPU/106 | 9.39 |
| | | | | | BRIS/87/HEPU/713 | 7.79 |
| | | | | | WB1B-M3 | 22.79 |
| | | | | | BRIS/83/HEPU/106-2ID$_{10}$ | 23.99 |
| | | | | | BRIS/87/HEPU/713-M3 | 42.33 |

na – not available; nd – not defined

[1)] For easy comparison, a conversion of µg/ml to µM was made in the case when the original dose was expressed in µg/ml. Converted data is highlighted in bold.

[2] concentration of drug which reduces the number of *G. lamblia* colonies to 50% of the control number

[3] minimum lethal concentration (MLC): the lowest concentration of the drug in which no *G. lamblia* colonies grew

[4] concentration of drug required to inhibit $^3$H-thymidine uptake by 50%

[5] Boreham et al. 1984

[6] adherence to Caco2 cells

[7] adherence to culture vessel walls

[8] drug concentration required to reduce the growth or adherence of treated cells to Caco2 cells by 50% ($ID_{50}$) as compared with untreated controls

[9] concentration of the drug that inhibited growth by 50% as calculated by probit analysis

[10] the lowest concentration of the drug at which no viable trophozoites were observed by subculture

[11] the lowest concentration of drug with which no viable organisms were observed at the termination of the assay

[12] ATP-based luminescence (corresponding to live cells/well) measured using a luminometer (BioTek), and transformed to a proportion of the negative control value. Prism software (GraphPad) was used to fit Hill plots to dose response data and to calculate IC50 values, via the "log (inhibitor) vs. normalized response—variable slope" module
